# Neuroanatomical variability associated with early substance use initiation: Results from the ABCD Study

**DOI:** 10.1101/2024.03.06.24303876

**Authors:** Alex P. Miller, David A. A. Baranger, Sarah E. Paul, Hugh Garavan, Scott Mackey, Susan F. Tapert, Kimberly H. LeBlanc, Arpana Agrawal, Ryan Bogdan

## Abstract

The extent to which neuroanatomical variability associated with substance involvement reflects pre-existing risk and/or consequences of substance exposure remains poorly understood. In the Adolescent Brain Cognitive Development^SM^ (ABCD®) Study, we identify associations between global and regional differences in brain structure and early substance use initiation (i.e., occurring <15 years of age; ns_analytic_=6,556-9,804), with evidence that associations precede initiation. Neurodevelopmental variability in brain structure may confer risk for substance involvement.

## Main

The widespread prevalence of substance use and its devastating consequences constitute a growing international public health concern.^1^ Extensive efforts are underway to identify markers of risk for substance involvement (e.g., initiation, escalating use, problematic use) and characterize the biological mechanisms through which it impacts health. Predominantly cross-sectional neuroimaging studies have reliably found that various forms of substance involvement are associated with smaller brain volumes and thinner cortex.^2–5^ While these associations are typically interpreted to reflect neurotoxic consequences of substance exposure, accumulating data also highlight that substance-related variability in brain structure may, at least partially, reflect pre-existing vulnerability.^6–10^

Using data (ns_analytic_=6,556-9,804) from the longitudinal Adolescent Brain Cognitive Development^SM^ (ABCD®) Study,^11^ we tested whether baseline brain structure (i.e., global and regional volume, cortical thickness, surface area, and sulcal depth; n_imaging-derived phenotypes_=297) is associated with early substance use initiation (i.e., occurring <15 years of age; **Online Methods**). Neuroimaging data from the initial study visit (baseline; 9-11 years of age) and substance use data from annual in-person (baseline; 1-, 2-, 3-year follow-up [FU]) and mid-year phone (6-, 18-, 30- month FU) assessments were used (**Online Methods; Table S1**). By the 3-year FU session, 3,460 participants reported using any substance with alcohol (n=3,123), nicotine (n=431), and cannabis (n=212) being most common; 6,344 participants reported not using any substances (**Figure S1**; **Table S2**). Given emerging evidence that neuroimaging correlates of substance involvement may partially reflect predispositional risk, we hypothesized that variability in baseline brain structure would predict early substance use initiation.

Linear mixed-effect models (**Online Methods**) revealed that eight imaging-derived phenotypes were associated with any early substance use initiation after Bonferroni correction (**Online Methods; Table S3-S4**). Any substance use initiation was associated with larger global brain structure indices (n=5) including whole brain and total intracranial, cortical, and subcortical volumes and greater total cortical surface area (βs=0.038-0.048, *P*s<6×10^-6^; **Figure 1**). Regionally (n=3), thinner right rostral middle frontal gyrus, thicker left lingual gyrus, and larger right lateral occipital gyrus volume were associated with any substance use initiation (|β|s=0.032-0.035, *P*s<2.65×10^-5^; **Figure 2A**; **Figure 3**). An additional 36 regional imaging-derived phenotypes were associated with any substance use initiation using less stringent false discovery rate (FDR) correction (**Online Methods; Figure 2A**; **Figure 3; Table S3**). Of all regional associations (n=39), the majority were with cortical thickness (56.4%). Notably, any substance use initiation was characterized by thinner cortex in all frontal regions (n=9), but thicker cortex in all other lobes (n_occipital_=6, n_parietal_=1, n_temporal_=6). Any substance use initiation was also associated with larger regional brain volumes (n_subcortical_=3, n_cortical_=7), deeper regional sulci (n=3), and differences in regional cortical surface area (n=4). Effect sizes were consistent with those observed in neuroimaging studies of other complex traits, highlighting small but potentially important effects.^12^

**Figure 1.**
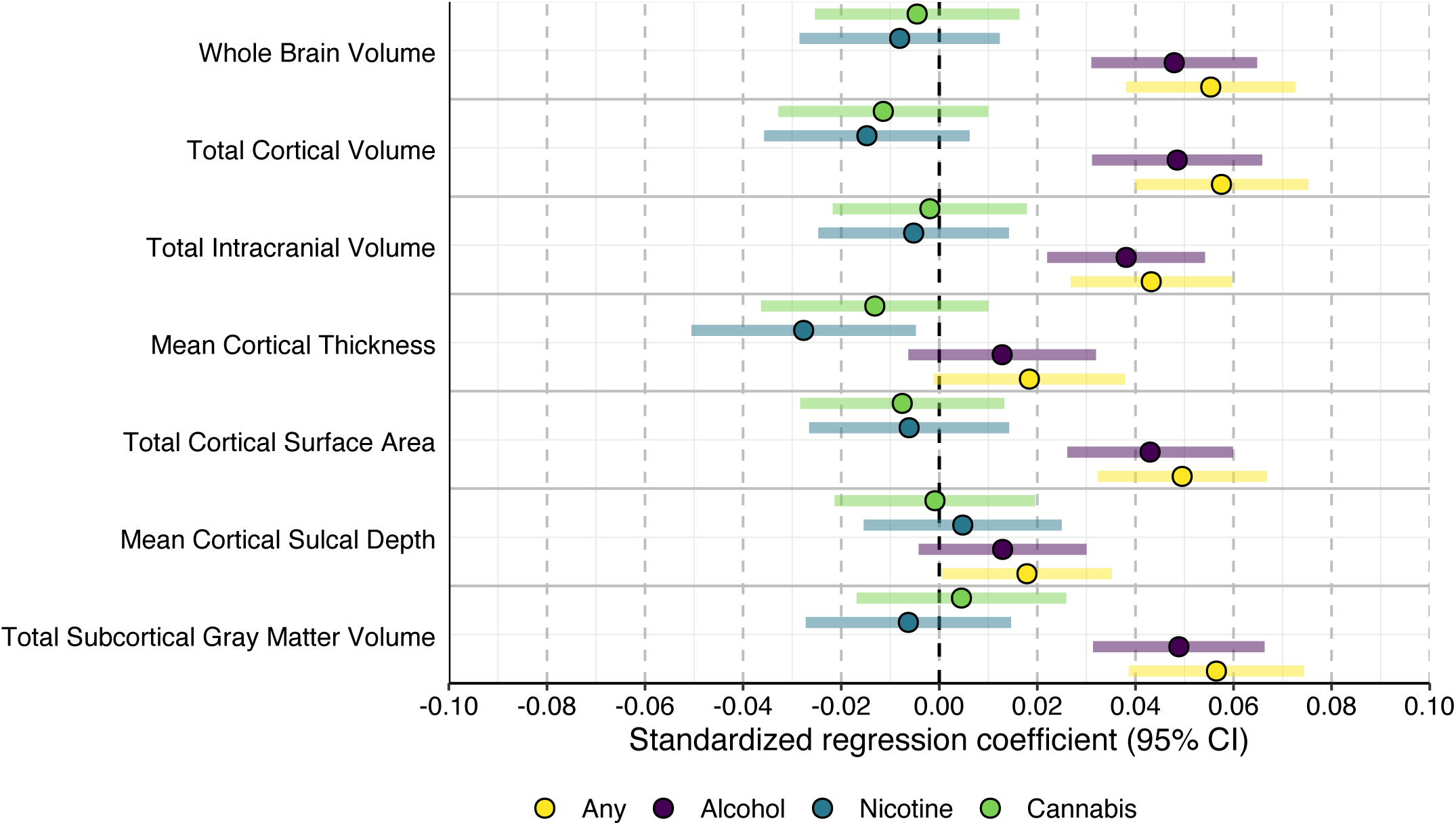
Global brain metric associations with lifetime use of substances in ABCD. Standardized regression coefficients with 95% confidence intervals for associations between global metrics and substance use initiation.

**Figure 2.**
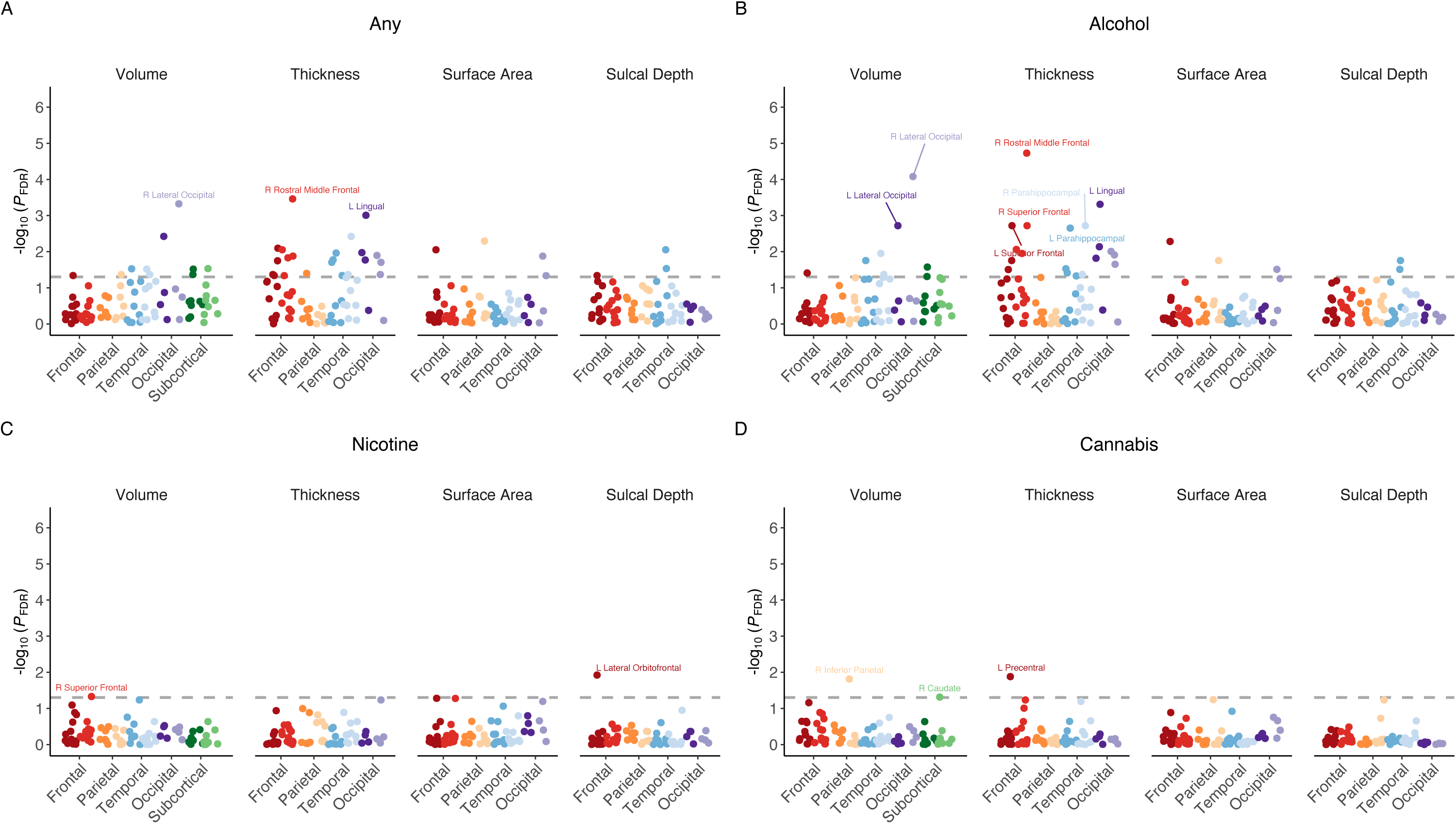
Regional brain structure associations with lifetime use of any substances (A), alcohol (B), nicotine (C), and cannabis (D) in ABCD. –log_10_-transformed FDR-corrected *P*-values (*P*_FDR_) from mixed-model regressions are plotted for all regional association analyses and grouped within neuroanatomical metrics for each substance (i.e., cortical and subcortical volume, thickness, surface area, and sulcal depth). *P*-values are aggregated and color coded by cortical lobes and subcortical regions with darker colors reflecting left hemisphere and lighter colors reflecting right hemisphere for each region (e.g., dark red = left frontal lobe; lighter red = right frontal lobe). Though often considered separate from frontal, parietal, and temporal lobes, and located at their junction, for simplicity the insular cortex is plotted here along with temporal regions. Dashed gray line reflects *P*_FDR_<.05. For any substance and alcohol use (**A-B**), labeled regions reflect associations that are Bonferroni-significant for all study comparisons (*P*<.05/1,188=4.21×10^-5^). For nicotine and cannabis use (**C-D**), labeled regions reflect FDR-significant associations.

**Figure 3.**
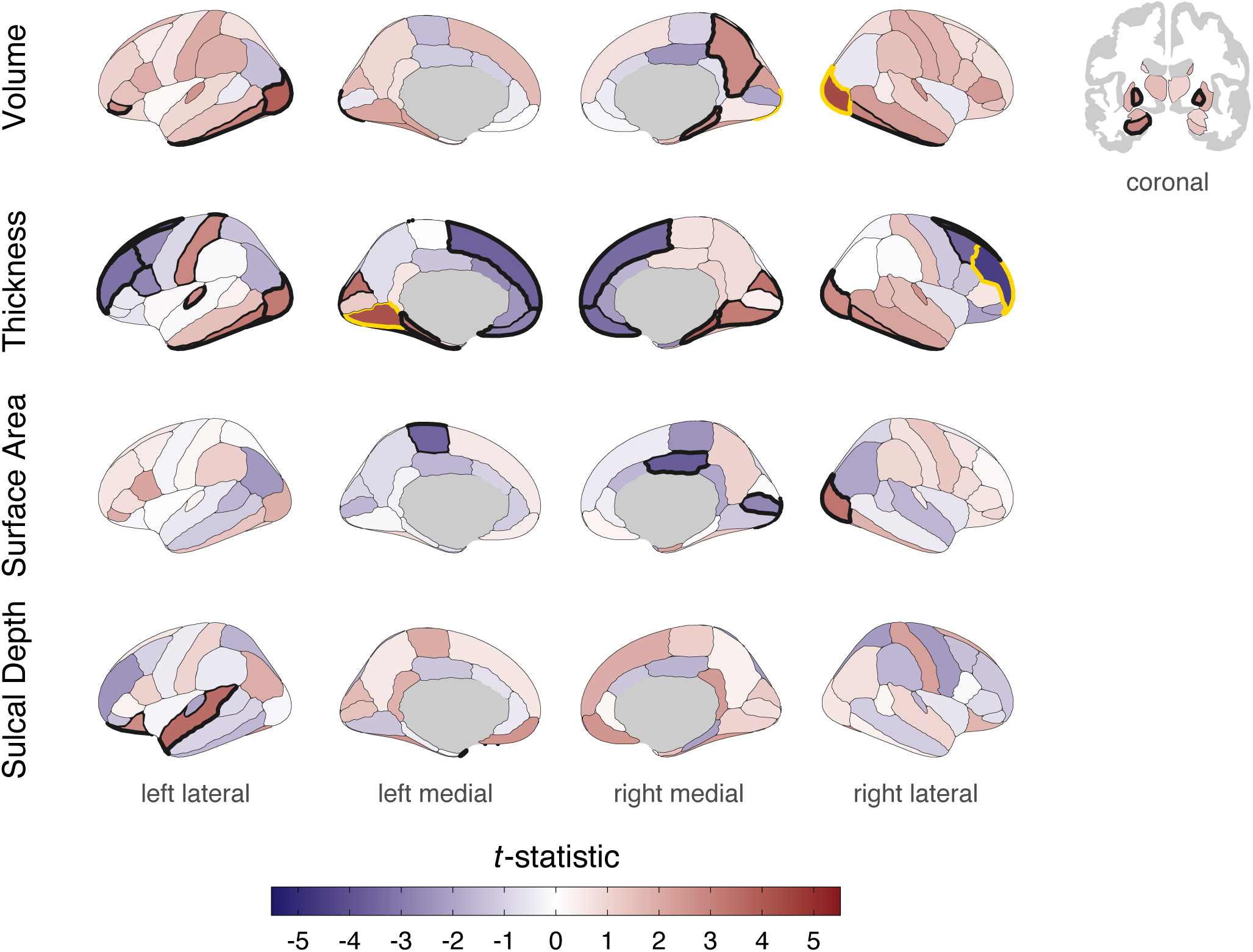
Regional cortical and subcortical associations with any lifetime substance use initiation in ABCD. Cortical and subcortical patterning of associations with substance use plotted as *t*-statistics (red = positive association, blue = negative association). Regions with bold outlines exhibit FDR-significant associations and those outlined in yellow are Bonferroni-significant for all study comparisons. Bonferroni-significant regions: volume of R lateral occipital; thickness of R rostral middle frontal and L lingual. FDR-significant regions: volume of R/L globus pallidus and inferior temporal, and R parahippocampal and precuneus, and L hippocampus, lateral occipital, and pars orbitalis; thickness of R/L cuneus, inferior temporal, lateral occipital, medial orbitofrontal, parahippocampal, and superior frontal, and R caudal middle frontal, frontal pole, and lingual, and L fusiform, pars opercularis, postcentral, rostral middle frontal, and transverse temporal; surface area of R lateral occipital, pericalcarine, and posterior cingulate, and L paracentral; sulcal depth of L superior temporal and temporal pole. See also **Table S3**.

Secondary analyses compared the three most commonly used substances (alcohol, nicotine, and cannabis) to no substance use (**Online Methods**). Unsurprisingly, given the preponderance of alcohol use initiation in this sample, alcohol findings largely recapitulated those observed for any substance use (**Supplementary Data**; **Figure 2B; Figure S2)**. Nicotine use was associated with lower right superior frontal gyrus volume and deeper left lateral orbitofrontal cortex sulci, and cannabis use was associated with thinner left precentral gyrus and lower right inferior parietal gyrus and caudate volumes following FDR correction; however, these associations were not robust to Bonferroni correction (**Supplementary Data**; **Figures 2C-D; Figures S3-S4; Table S7-10**). *Post hoc* analyses revealed that all FDR- and Bonferroni-significant associations remained significant (*P*_FDR_<.05) when including prenatal exposure as a covariate. Further, several Bonferroni-significant associations remained significant when removing participants who endorsed substance initiation prior to the baseline neuroimaging session, including most global (e.g., whole brain) and subcortical (e.g., globus pallidus) volume associations and many cortical thickness findings such as the right rostral middle frontal gyrus for alcohol (**Online Methods; Supplementary Data**; **Table S11**). Effect sizes in the full sample were highly correlated with those from the sample that excluded baseline initiation (*r*s=0.60-0.94; **Supplementary Data**; **Figure S5**).

Early substance use initiation is associated with escalating use, use of multiple substances, and development of substance use disorders and other adverse life outcomes (e.g., reduced educational attainment, elevated rates of non-substance psychopathology).^13–16^ We identified global and regional brain structure correlates of late childhood and early adolescent substance use initiation (any, alcohol, nicotine, and cannabis), many of which are evident prior to any substance exposure. The direction of association between cortical thickness and substance use initiation was regionally specific; the cortical mantle was thinner in the prefrontal cortex, but thicker in temporal, occipital, and parietal regions among youth who initiated substance use. While age is associated with cortical thickness broadly across brain regions,^17^ our data suggest that regionally-specific differences in brain structure development during childhood, rather than general age-related trends, may confer vulnerability to substance use initiation. Substance use disorders and in particular alcohol use disorder have been characterized by broad reductions in cortical thickness in multiple brain regions with largest effects found in the frontal cortex.^2,3^ Our observation that regional associations precede substance use initiation, including the finding that reduced cortical thickness in the right rostral middle frontal gyrus precedes alcohol use initiation and that effect sizes were highly similar when excluding participants with baseline initiation, challenges predominant interpretations that these associations arise due to neurotoxic consequences of substance exposure^2,3,18^ and increases the plausibility that these frontal correlates may, at least partially, reflect markers of predispositional risk.^10,19^

This finding has important implications for brain-based theoretical models of addiction. The stage-based neurobiological model of addiction speculates that predominantly substance-induced variability in frontal regions contributes to later stages of addiction wherein compulsive use and craving develop following repeated-drug pairings and related disruption of prefrontal afferent regulation of subcortical reward and stress-related circuitry.^20^ These changes may make it difficult for individuals to stop using, even if there are deep subjective desires to do so.^20^ Our findings suggest that structural differences in the prefrontal cortex may predispositionally contribute to *initial* stages of substance involvement. Thinner frontal cortex alongside larger subcortical volumes comports with neurodevelopmental theories suggesting that typical asynchronous regional brain maturation (i.e., rapid subcortical development and later prefrontal development) leaves adolescents vulnerable to substance involvement by promoting emotional saliency in the context of underdeveloped cognitive control.^21,22^ It is notable that total cortical thickness peaks at 1.7 years of age and steadily declines throughout life with limited evidence of regionally-specific trajectories.^23^ On the other hand, subcortical volumes peak at 14.4 years of age and generally remain stable before steep declines in later life.^23^ Large-scale studies tracking neurodevelopment from infancy to early childhood and later substance involvement are necessary to parse whether substance-related brain structure differences may be attributable to differential early maturation and/or accelerated decline in later childhood. Such questions may be most readily addressed by integrating across cohort datasets such as the Healthy Brain and Child Development (HBCD)^24^ and ABCD studies.

Unlike negative associations between whole and regional brain volumes with adult substance use and substance use disorders,^3–5^ we found that greater global/regional cortical and subcortical volumes and total cortical surface area were associated with substance use initiation. Notably, larger globus pallidus volume, here associated with substance use initiation and in other ABCD studies associated with sensation seeking,^25^ has been linked to both occasional use and substance use disorders in adults,^2^ highlighting a plausibly stable risk feature from precocious use and experimentation to later problematic use. Moreover, greater cortical surface area has previously been observed to be correlated with genetic risk for alcohol use and sensation seeking in independent samples,^26,27^ as well as with family history of substance problems in this sample,^28^ supporting the interpretation that greater surface area may reflect the influence of pre-existing risk factors. Importantly, our findings here are not incompatible with subsequent declines in brain volumes that may arise from substance exposure and/or differences in later brain development conferring risk for substance involvement progression.

While our study is the first of variability in brain structure preceding substance involvement sufficiently powered to detect expected small effects for any substance and alcohol use initiation,^12^ we had limited power to detect anticipated small effects for less frequently endorsed individual substances (i.e., 80% power to detect |β|>0.042 at *P*<.005 for cannabis or nicotine). It will be important for future work to examine potential sources of variability that contribute to brain structure correlates of substance use initiation (e.g., genomic, prior environmental exposures). Given the large heritability of brain structure^29^ and evidence of family history of substance use problems as a risk factor for thinner frontal cortices,^30,31^ genetically-informed studies are needed to parse the molecular substrates of this variability and evaluate the plausibility of environmental causality including through possible socioeconomic differences.

Limitations notwithstanding, our study identifies brain structure correlates of substance use initiation that are present prior to substance exposure. Alongside evidence from genetically-informed designs (e.g., discordant twin and sibling),^8,9,32^ our longitudinal data increasingly place interpretations that substance-related variability solely arises as a result of substance exposure on a procrustean bed.

## Data availability

Data used in the preparation of this article were obtained from the Adolescent Brain Cognitive Development^SM^ (ABCD) Study (https://abcdstudy.org), held in the NIMH Data Archive (NDA). This is a multisite, longitudinal study designed to recruit more than 10,000 children aged 9-10 and follow them over 10 years into early adulthood. The ABCD Study® is supported by the National Institutes of Health and additional federal partners under award numbers U01DA041048, U01DA050989, U01DA051016, U01DA041022, U01DA051018, U01DA051037, U01DA050987, U01DA041174, U01DA041106, U01DA041117, U01DA041028, U01DA041134, U01DA050988, U01DA051039, U01DA041156, U01DA041025, U01DA041120, U01DA051038, U01DA041148, U01DA041093, U01DA041089, U24DA041123, U24DA041147. A full list of supporters is available at https://abcdstudy.org/federal-partners.html. A listing of participating sites and a complete listing of the study investigators can be found at https://abcdstudy.org/consortium_members/. ABCD consortium investigators designed and implemented the study and/or provided data but did not necessarily participate in the analysis or writing of this report. This manuscript reflects the views of the authors and may not reflect the opinions or views of the NIH or ABCD consortium investigators. The ABCD data repository grows and changes over time. The ABCD data used in this report came from http://dx.doi.org/10.15154/1520591 (ABCD Annual Release 3.0) and http://dx.doi.org/10.15154/8873-zj65 (ABCD Annual Release 5.0). DOIs can be found at https://nda.nih.gov/abcd/abcd-annual-releases.html.

## Acknowledgements

This study was supported by R01DA54750 (RB, AA). Additional funding included: APM (T32DA015035), DAAB (K99AA030808), SEP (F31AA029934), AA (R01DA54750), RB (R01DA54750, R21AA027827, U01DA055367). Data for this study were provided by the Adolescent Brain Cognitive Development (ABCD) study which was funded by awards U01DA041022, U01DA041025, U01DA041028, U01DA041048, U01DA041089, U01DA041093, U01DA041106, U01DA041117, U01DA041120, U01DA041134, U01DA041148, U01DA041156, U01DA041174, U24DA041123, and U24DA041147 from the NIH and additional federal partners (https://abcdstudy.org/federal-partners.html).

## Online Methods

### Participants

The Adolescent Brain Cognitive Development^SM^ (ABCD) Study^11^ is a longitudinal study of complex behavior and biological development from middle childhood to late adolescence/young adulthood. Children (n=11,875), aged 8.9-11 years of age at baseline (born between 2005-2009), were recruited from 22 research sites across the United States. Parents/caregivers provided written informed consent, and children assent, to a research protocol approved by the institutional review board at each site (https://abcdstudy.org/sites/abcd-sites.html). For the present analyses, baseline neuroimaging data were drawn from ABCD data release 3.0, while additional covariate variable data and substance use assessment data were drawn from ABCD data release 5.0. After excluding participants with missing substance use initiation, structural MRI (sMRI), and relevant covariate data described below, final analytic samples ranged from 6,556-9,804 participants.

### Substance Use Initiation

Substance use data were obtained from annual in-person (i.e., baseline and follow-ups at 1-year [FU1], 2-year [FU2], and 3-year [FU3]) and mid-year phone (i.e., 6-month [FU0.5], 18-month [FU1.5], 30-month [FU2.5]) substance use interviews (data release 5.0). Participants endorsing substance use at any point from baseline to FU3 (i.e., lifetime use) were included in substance use initiation groups. Alcohol (n=808) and nicotine (n=34) use endorsed solely in the context of religious ceremonies were coded as missing to restrict comparisons to childhood and adolescent use outside of these settings.

***Alcohol use initiation*** was defined as endorsement of ‘sipping’ or ‘full drinks’ of alcohol from baseline to FU3. ***Nicotine use initiation*** was defined as use of nicotine products in any form or route of administration, including puffing or smoking tobacco cigarettes or cigars, e-cigarettes, hookah or pipes, or using smokeless tobacco or chew, or nicotine patches, from baseline to FU3. ***Cannabis use initiation*** was defined as use of cannabis in any form or route of administration, including puffing or smoking or vaping flower, oils, or concentrates, smoking blunts, or consuming edibles or tinctures, but not including synthetic cannabis or cannabis-infused alcoholic drinks, which were included under any substance use, from baseline to FU3. ***Any substance use initiation*** was defined as alcohol, nicotine, or cannabis use initiation or use of any other illicit substance (e.g., hallucinogens, or misuse of prescription medications like stimulants or sedatives) from baseline to FU3. ***Substance naïve participants*** endorsed no use of any substance from baseline to FU3 while also having non-missing data for FU3 to protect against misclassification of participants who may have been substance naïve from baseline to FU2, but missing data for FU3, which could be indicative of onset of use. While more specific indices of quantity, frequency, and level of use are available for individual substances in these data, especially alcohol (e.g., ‘sips’ *vs.* ‘full drinks’, number of sipping occasions^12^), here we focused on broad-spectrum use *vs.* no use given currently low base rates of even initiation endorsed by children and early adolescent ABCD participants and expected small effects,^33,34^ and to provide more consistent variable definitions across substances. Additionally, prior work has shown that alcohol sipping at these ages in ABCD is consistently associated with impulsivity and other aspects of externalizing psychopathology, highlighting its relevance as an appropriate alcohol-related phenotype for children and early adolescents.^35^ For details on individual ABCD 5.0 release variables used to define these groups see **Table S2**.

### Structural Magnetic Resonance Imaging

Valid baseline imaging data were available for 11,556 out of 11,875 participants. Briefly, 1mm isotropic T1-weighted structural images were obtained via 3T MRI scanners (Siemens, Phillips, and GE) using either a 32- or 64-channel head-and-neck coil and completed T1-weighted and T2-weighted structural scans (1mm isotropic).^35^ sMRI scan protocols were harmonized across the three MRI vendor platforms to minimize variability. Real-time motion detection and correction was implemented to mitigate the influence of head motion. sMRI processing was completed using FreeSurfer version 5.3.0 through standardized processing pipelines and described quality control (QC) procedures performed by the ABCD Study^®^ Data Analysis, Informatics and Resources Center using the FreeSurfer image analysis suite (http://surfer.nmr.mgh.harvard.edu/).^36^ Participants who did not pass FreeSurfer QC measures (i.e., at least one T1 scan that passed all QC metrics) were excluded from analyses. Cortical reconstruction and volumetric segmentation was performed by the ABCD Study^®^ Data Analysis, Informatics and Resources Center using the FreeSurfer image analysis suite (http://surfer.nmr.mgh.harvard.edu/). This pre-processing includes removal of non-brain tissue using a hybrid watershed/surface deformation procedure,^36^ automated Talairach transformation, segmentation of the subcortical white matter and deep gray matter volumetric structures, intensity normalization, tessellation of the gray/white matter boundary, automated topology correction, and surface deformation following intensity gradients.^36^ Cortical images were registered to the Desikan atlas, which was based on individual cortical folding patterns to match cortical geometry across participants. The cerebral cortex was parcellated into 34 regions per hemisphere based on the gyral and sulcal structure. For sulcal depth, regions that moved outward during inflation were positive and represent the depths of sulci, and regions that moved inward were negative and represent the height of gyri.^38^

For the current analyses, cortical gray matter volume and total cerebral white matter volume aligned to the Desikan-Killiany atlas were extracted from ABCD data release 3.0. Only sMRI data that passed QC tests were retained. In total, 297 imaging-derived phenotypes (IDPs) were examined in the current study. These included seven global IDPs (i.e., whole brain volume, total intracranial volume, total cortical volume, total cortical surface area, mean cortical thickness, mean cortical sulcal depth, and total subcortical gray matter volume); bilateral volume of nine subcortical structures (i.e., accumbens, amygdala, caudate, cerebellar cortex, cerebellar white matter, globus pallidus, hippocampus, putamen, and thalamus; total *k*=18); and total volume, mean thickness, total surface area, and mean sulcal depth of 34 bilateral cortical regions according to the Desikan-Killiany atlas (total *k*=68 per metric).

### Statistical Analysis

All variables were *z*-scored prior to analyses. The ‘lme4’ R (v4.2.1) package (v1.1-30)^41,42^ was used to conduct a series of independent linear mixed effect regressions wherein IDPs were regressed on substance use initiation groups. Primary analyses contrasted any substance use initiation (n=3,460) vs. no initiation (n=6,344); secondary analyses considered alcohol (n=3,123), nicotine (n=431), and cannabis (n=212) use initiation independently and contrasted them against no substance initiation (n=6,344).Variables known to be associated with substance use initiation, brain structure, or both were included as fixed effect covariates in all analyses: baseline age and baseline age-squared,^43,44^ sex,^45–47^ pubertal status,^29,48^ familial relationship (i.e., sibling, twin, triplet),^49,50^ and MRI scanner model.^51^ For pubertal status, parents and children both completed a 5-item scale on the child’s pubertal development,^52,53^ combined to a summary score. The parent rating was used as the primary measure. Given high congruence between parent and child ratings reported previously,^54,55^ the child rating (n=223) was used if the parent rating was unavailable to reduce data loss to missingness as done in previous ABCD studies.^56–61^ Sociodemographic variables, indexing socioeconomic status and social determinants of health, are also associated with both brain structure development and substance use outcomes.^12^ These variables were not included as covariates as they may plausibly influence observed associations in meaningful ways, in which case exploring the mechanisms by which they might influence associations, a research question beyond the scope of the current study, would be more appropriate than controlling for them. For models examining associations with regional and structural IDPs, global metrics were also included as covariates: total intracranial volume for regional cortical and subcortical structural volumes, total cortical surface area for regional surface area, mean cortical thickness for regional thickness, and mean sulcal depth for regional sulcal depth. Random intercepts were modeled for effects of family nested within data collection sites.

Multiple testing corrections were applied using a 5% false discovery rate (FDR) first across all tests examining associations between IDPs and any substance use initiation (*n*=297 tests). A separate FDR correction was applied to all tests collectively examining associations between IDPs and alcohol, nicotine, and cannabis use initiation (*n*=891 tests). A Bonferroni correction was also applied to all tests conducted as part of the study (*P*=.05/1,188=4.21×10^-5^) to prioritize especially robust results. Collectively, our method of multiple testing correction, employing both FDR- and Bonferroni-corrections, was selected based on expected small effect sizes^62^ and to balance the risk for false positive results (Bonferroni-correction), which may be less likely to replicate or generalize to other samples, and emphasize more robust associations, and the risk for false negative results that comes with correcting for a large number of tests (FDR-correction).

### Post Hoc Analyses

Given findings from our recent work and work by others in ABCD demonstrating potential effects of prenatal substance exposure on both substance use initiation^55,63^ and childhood brain structure,^64^ *post hoc* analyses examined possible effects of prenatal substance exposures on FDR-significant associations. Baseline retrospective report of maternal substance use before or after knowledge of pregnancy, either specific to alcohol, nicotine, or cannabis, or any substance including other substances (i.e., cocaine, opioids, methamphetamine, or other drugs), was added to models as a fixed effect covariate. Significant effects following a second FDR-correction (*P*_FDR_<.05) for IDPs in these follow-up analyses were interpreted to reflect associations independent of prenatal substance exposure. Moreover, as a significant portion of the sample initiated substance use prior to the baseline study session (n=2,257), additional post hoc analyses examined whether FDR-significant associations remained when restricting substance use initiation groups to only participants endorsing initiation following the baseline assessment (any n=1,203; alcohol n=933; tobacco n=322; cannabis n=201) to test whether IDP correlates are present prior to initiation; here significant associations following a second FDR-correction (*P*_FDR_<.05) were interpreted to reflect plausible evidence that these associations may reflect predispositional vulnerability. Finally, both restriction of analytic samples to post-baseline substance use initiation and inclusion of prenatal exposure as a covariate were incorporated in *post hoc* tests to examine effects which might reflect predispositional variability independent of prenatal substance exposure.

### Regional Brain Plots

Regional brain plots were constructed using the *ggseg* package in R.^65^ Cortical and subcortical patterning of associations with substance use were plotted as *t*-statistics (red = positive association, blue = negative association) according to the Desikan–Killiany cortical atlas^66^ and the Automatic Segmentation of Subcortical Structures atlas^66^, respectively.

## Notes

### Competing Interest Statement

The authors have declared no competing interest.

