## Supplementary Data for "Neuroanatomical variability associated with early substance use initiation: Results from the ABCD Study"

Alex P. Miller, Ph.D. [1], David A. A. Baranger, Ph.D. [2], Sarah E. Paul, M.A. [2], Hugh Garavan, Ph.D. [3], Scott Mackey, Ph.D. [3], Susan F. Tapert, Ph.D. [4], Kimberly H. LeBlanc, Ph.D. [5], Arpana Agrawal, Ph.D. [1], & Ryan Bogdan, Ph.D. [2]

[1] Department of Psychiatry, School of Medicine, Washington University in St. Louis, St. Louis, MO, United States; [2] Department of Psychological and Brain Sciences, Washington University in St. Louis, St. Louis, MO, United States; [3] Department of Psychiatry, University of Vermont Lamer College of Medicine, Burlington, VT, United States; [4] Department of Psychiatry, University of California San Diego, San Diego, CA, United States; [5] Division of Extramural Research, National Institute on Drug Abuse, Bethesda, MA, United States

**Table of Contents**

**Substance Use Initiation.**
Patterns of substance use initiation in the current analytic sample ...………………………Page 1

**Brain Structure Correlates.**
Expanded presentation of association results for alcohol, nicotine, and cannabis use …….. Page 2

***Post Hoc* Results.**
Expanded presentation of associations results from *post hoc* analyses ……………………. Page 3

**Comparison of Effect Sizes for Full *vs.* Restricted Analytic Samples** ………………….Page 4

**References** ……………………………………………………………………….Page 5

**Figure S1.**
Alcohol, nicotine, and cannabis use in current analytic ABCD
sample through follow-up year 3 ……………………………………………………... Page 6

**Figure S2.**
Regional cortical and subcortical associations with
lifetime alcohol use initiation in ABCD ………………………………………………… Page 7

**Figure S3.**
Regional cortical and subcortical associations with
lifetime nicotine use initiation in ABCD ……………………………………………….. Page 8

**Figure S4.**
Regional cortical and subcortical associations with
lifetime cannabis use initiation in ABCD ………………………………………………. Page 9

**Figure S5.**
Correlations between effect sizes in analyses in the full sample and analyses
restricted to participants without baseline substance use initiation ………………………. Page 10

### Substance Use Initiation

A total of 3,460 (35.3%) ABCD participants in the current analytic sample endorsed lifetime use of substances outside the context of religious ceremonies during at least one assessment, while 6,344 (64.7%) participants reported no substance use (substance naïve). The vast majority of those endorsing substance use initiation by FU3 had reported using alcohol ($n$=3,123; 90.2%) with fewer participants reporting use of nicotine ($n$=431; 12.5%) and cannabis ($n$=212; 6.1%), respectively (**Table S2**). Moreover, nearly all participants with lifetime alcohol use reported having 'sips' of alcohol ($n$=2,931; 94.2%) without endorsing 'full drinks' ($n$=192; 5.8%). Reported routes of administration of nicotine and cannabis varied across participants and assessments. In general, however, e-cigarettes or vaping were the predominant method of nicotine use with 76.5% of those reporting specific routes of administration endorsing e-cigarette use during at least one assessment and 61.5% endorsing e-cigarette use exclusively. Tobacco cigarette use was endorsed by 18.7% of those reporting specific routes of administration, while a similar proportion (22.5%) endorsed other forms of nicotine use (i.e., cigars, hookahs, pipes, chew, nicotine replacement patches). For cannabis, the most frequently endorsed specific route of administration was smoking flower (65.2%), followed by consuming edibles (48.9%), and using oils, concentrates, or tinctures (37.1%). Only 23.0% of participants endorsing specific routes of cannabis administration reported vaping flower, concentrates, or oils. Two-hundred and thirteen participants endorsed use of substances other than alcohol, nicotine, or cannabis (e.g., hallucinogens, stimulants, sedatives; **Table S2** notes) with more than 50% of these participants endorsing use of only these other substances.

Notably, there was considerable overlap among participants reporting use of alcohol, nicotine, and cannabis ($\chi^2(3)$=251.35, $P$=3.33×$10^{-54}$; **Figure S1A**). For instance, more than 65%

of those endorsing nicotine or cannabis use also endorsed use of the other substance and/or alcohol, and 68 participants endorsed use of all three. As would be expected, cumulative lifetime endorsement of alcohol, nicotine, and cannabis use increased from baseline to FU3 (**Figure S1B**). However, while smaller subsets of participants endorsed use of nicotine (*n*=109) or cannabis (*n*=11) during the initial baseline assessment, more than 70% of those endorsing use of alcohol thus far in the study had done so at baseline (*n*=2,190; **Table S2**). Increasing rates of endorsement of cannabis (+1,927%) and nicotine (+395%) use initiation from baseline to FU3 outpaced increasing prevalence in alcohol use initiation (+147%; **Figure S1B**). Notably, rates of prenatal substance exposure were consistently higher among participants in substance use initiation groups compared to substance naïve participants with some evidence of substance specificity (i.e., alcohol, nicotine, and cannabis use initiation groups demonstrated the highest rates of prenatal alcohol, nicotine, and cannabis exposure, respectively; **Table S2**).

**Brain Structure Correlates**

***Alcohol Use Initiation.*** There were 37 FDR-significant IDP associations with alcohol use initiation compared to substance naïvety. Thirteen of these remained significant after Bonferroni correction for all study tests conducted (**Table S5**; **Figure 2B**). In addition to identifying the same global and regional IDPs that were associated with any substance initiation (*n*=8) described above, the following additional IDPs (*n*=5) were also significant after Bonferroni correction: greater left lateral occipital volume and bilateral parahippocampal gyri cortical thickness and reduced bilateral superior frontal gyri cortical thickness (all $|\beta|$s>0.027, all $P\text{s} \leq 4 \times 10^{-5}$; **Figure S2**). As was the case for the any substance use initiation contrast, FDR-significant regional results were primarily for cortical thickness and reflected reduced thickness in the frontal cortex but greater thickness in all other areas. Similarly, there was evidence of greater left globus pallidus and hippocampal volume among alcohol use initiators (**Table S5; Figure S2**)**.** See **Table S6** for full association results with alcohol use initiation.

***Nicotine Use Initiation.*** Nicotine initiation was associated with two IDPs following FDR-correction: reduced volume of the right superior frontal gyrus (β=-0.028, $P$=2.06×10$^{-3}$) and greater mean sulcal depth of the left lateral orbitofrontal cortex (β=0.045, $P$=2.68×10$^{-4}$; **Figure 3C; Figure S3; Table S7**). These associations were not significant following Bonferroni adjustment for all tests. **Table S8** presents full results.

***Cannabis Use Initiation.*** Cannabis use initiation was significantly associated with reduced cortical thickness in the left precentral gyrus (β=-0.030, $P$=3.26×10$^{-4}$) and reduced volume of the right inferior parietal gyrus (β=-0.034, $P$=4.13×10$^{-4}$) and right caudate (β=-0.030, $P$=2.24×10$^{-3}$; **Figure 3D; Figure S4; Table S9**) following FDR correction; none of these associations were robust to Bonferroni correction for all tests. **Table S10** presents full results.

***Post Hoc* Results**

All FDR- and Bonferroni-significant associations remained significant after a second FDR-correction ($P_{FDR}$<.05) when covarying for separate substance-specific prenatal exposures or any prenatal substance exposure. After restricting analytic samples to participants reporting substance use initiation only after study entry (i.e., following the baseline session), 50% and 76.9% of Bonferroni-significant results for any substance and alcohol use initiation remained significant ($P_{FDR}$<.05), respectively (see **Tables S3 and S5**). Of FDR-significant associations, 20.5% and 45.9% remained significant ($P_{FDR}$<.05). All FDR-significant results for nicotine and cannabis use initiation remained significant ($P_{FDR}$<.05) when excluding participants endorsing baseline nicotine and cannabis use (**Tables S7 and S9**). When including prenatal substance exposure (any or substance-specific) as a covariate after restricting analytic samples to post-baseline initiation,

>22.7% of FDR-significant associations and >46.1% of Bonferroni-significant associations remained nominally significant. See **Tables S3, S5, S7, and S9** for full association results of *post hoc* tests and **Table S11** for a summary of all *post hoc* analyses.

**Comparison of Effect Sizes for Full *vs.* Restricted Analytic Samples**

Global and regional effect sizes were correlated between analyses that included all participants and analyses that were restricted to only participants with no substance use initiation at the baseline visit (**Figure S5**). As spatial auto-correlation prevents deriving a *P*-value directly from the correlation coefficient,[1] exchangeability block permutations with accelerated *P*-values were used to determine the significance of the correlations.[2,3] Analyses were restricted to unrelated participants in order to facilitate the identification of valid permutations of substance initiation variables that did not violate the hierarchical structure of the data (i.e., observations nested within families, nested within sites). All regression analyses were then re-run for 10,000 permutations of the substance initiation variables, and analyses then computed the correlation between effect sizes of the permuted data with the true effect sizes from analyses restricted to participants with no baseline substance use initiation. This then yields, for each correlation, 10,000 coefficients drawn from a valid null distribution (i.e., an empirical null distribution). A beta distribution was then fit to the empirical null distribution, and a *P*-value for each true correlation was computed.[4]

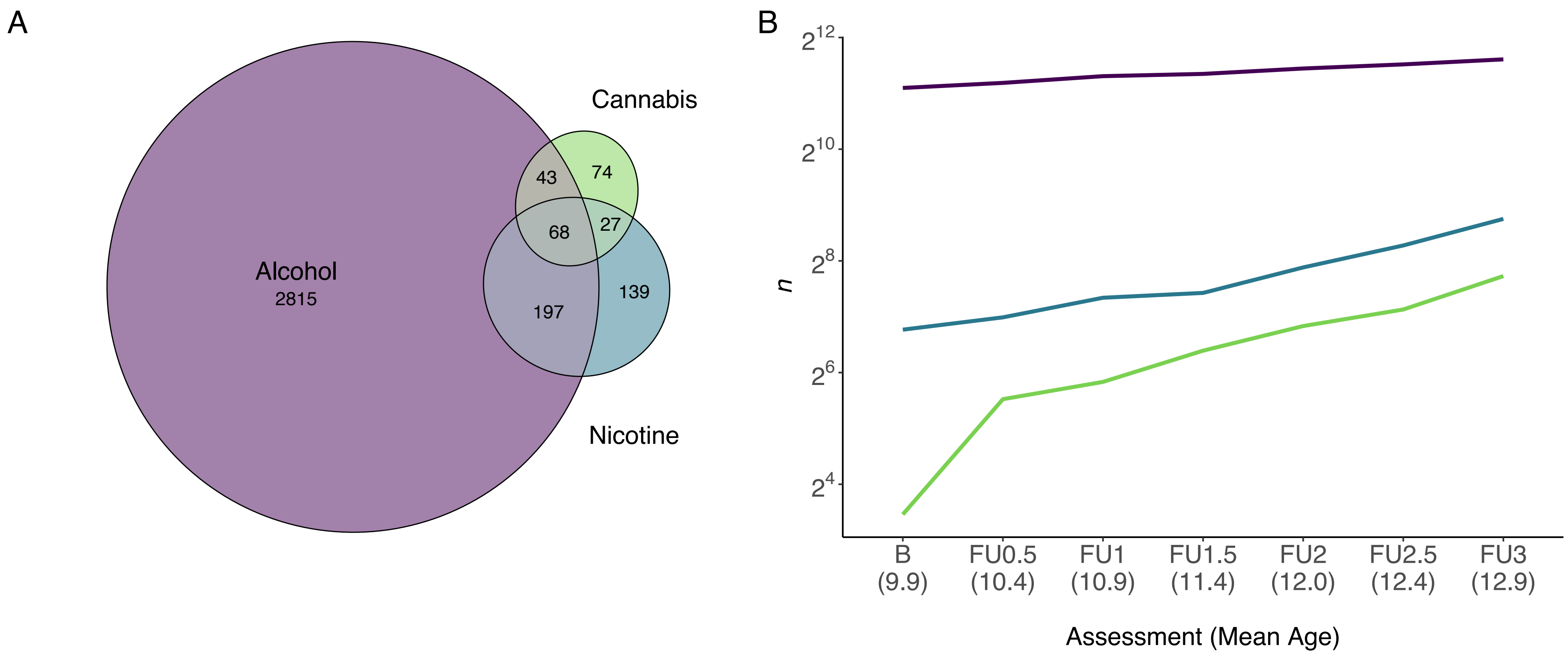


**Figure S1. Alcohol, nicotine, and cannabis use in current analytic ABCD sample through follow-up year 3. A.** Venn diagram exhibiting overlap amongst participants endorsing lifetime alcohol (including both 'sipping' and 'full drinks'), nicotine, and cannabis use. **B.** Cumulative lifetime alcohol (including both 'sipping' and 'full drinks'), nicotine, and cannabis use initiation from baseline ($M_{age}$=9.9 years) to third follow-up assessment ($M_{age}$=12.9 years). Y-axis log2-transformed sample size.

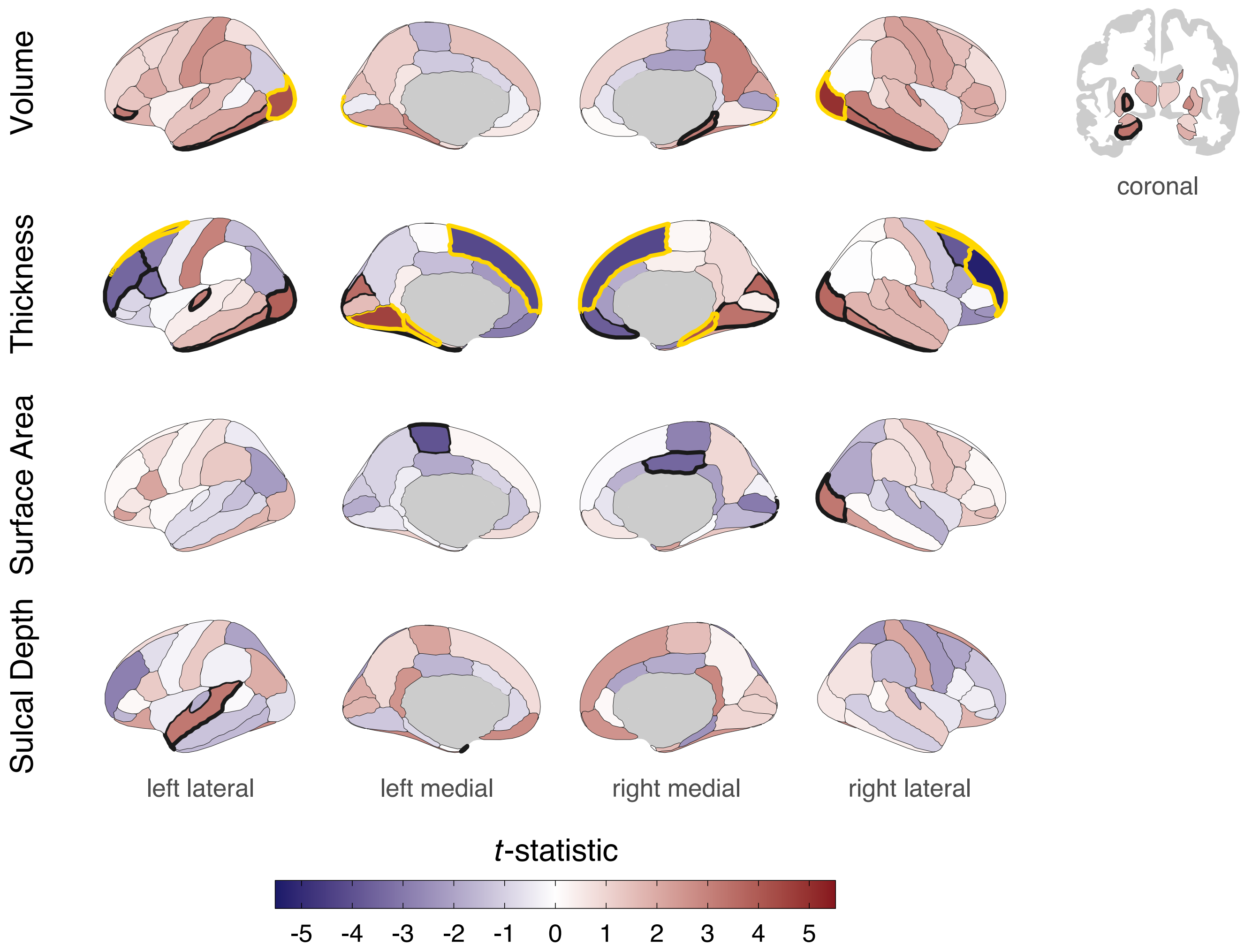


**Figure S2. Regional cortical and subcortical associations with lifetime alcohol use initiation in ABCD.** Cortical and subcortical patterning of associations with substance use plotted as *t*-statistics (red = positive association, blue = negative association). Regions with bold outlines exhibit FDR-significant associations and those outlined in yellow are Bonferroni-significant for all study comparisons. Bonferroni-significant regions: volume of R/L lateral occipital; thickness of R/L parahippocampal, R/L superior frontal, R rostral middle frontal, and L lingual. FDR-significant regions: volume of R/L inferior temporal, R parahippocampal, and L globus pallidus, hippocampus, and pars orbitalis; thickness of R/L cuneus, inferior temporal, and lateral occipital, R caudal middle frontal, lingual, and medial orbitofrontal, and L fusiform, pars opercularis, rostral middle frontal, and transverse temporal; surface area of R lateral occipital and posterior cingulate, and L paracentral; sulcal depth of L superior temporal and temporal pole. See also **Table S5.**

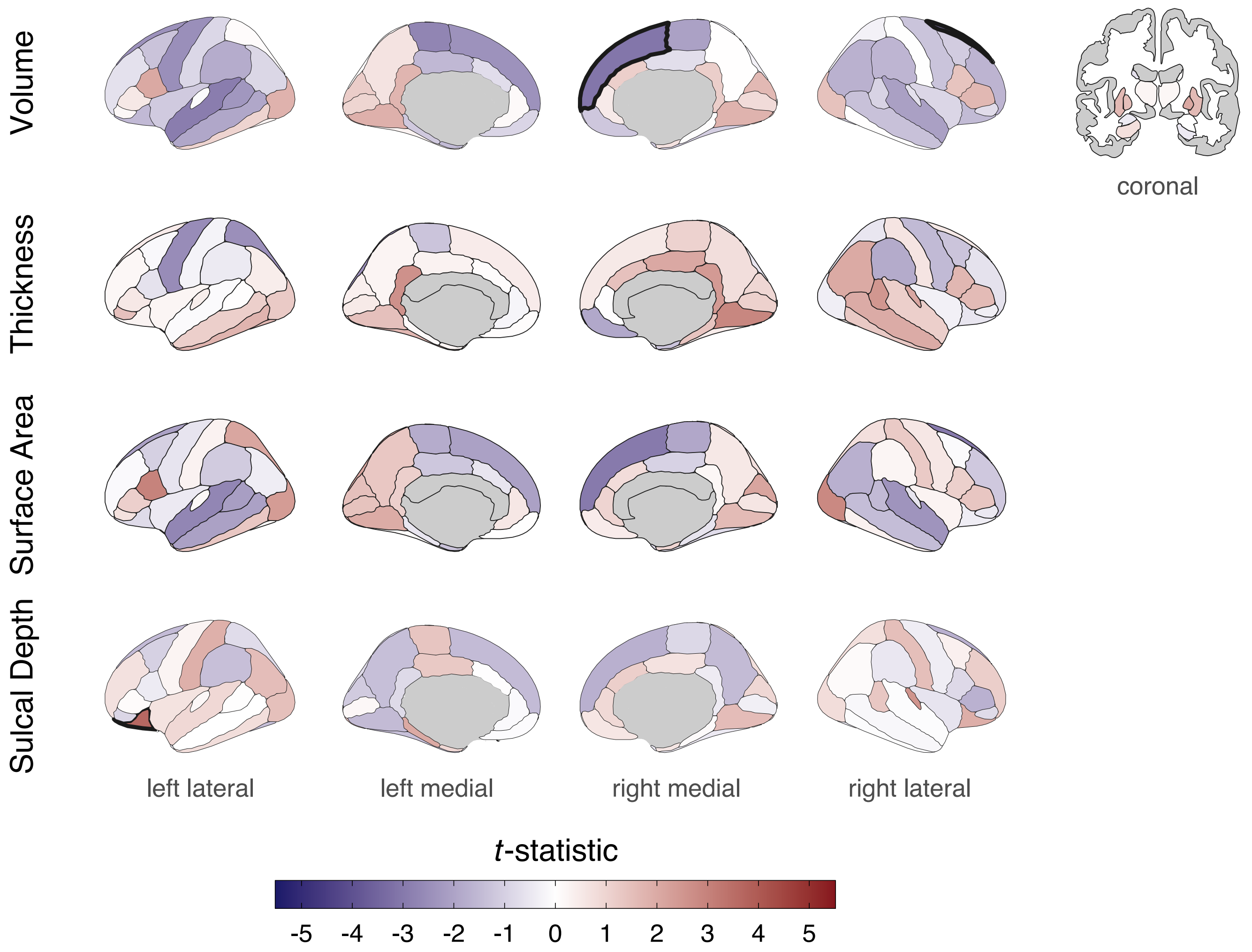


**Figure S3. Regional cortical and subcortical associations with lifetime nicotine use initiation in ABCD.** Cortical and subcortical patterning of associations with substance use plotted as *t*-statistics (red = positive association, blue = negative association). Regions with bold outlines exhibit FDR-significant associations: volume of R superior frontal and sulcal depth of L lateral orbitofrontal. See also **Table S7.**

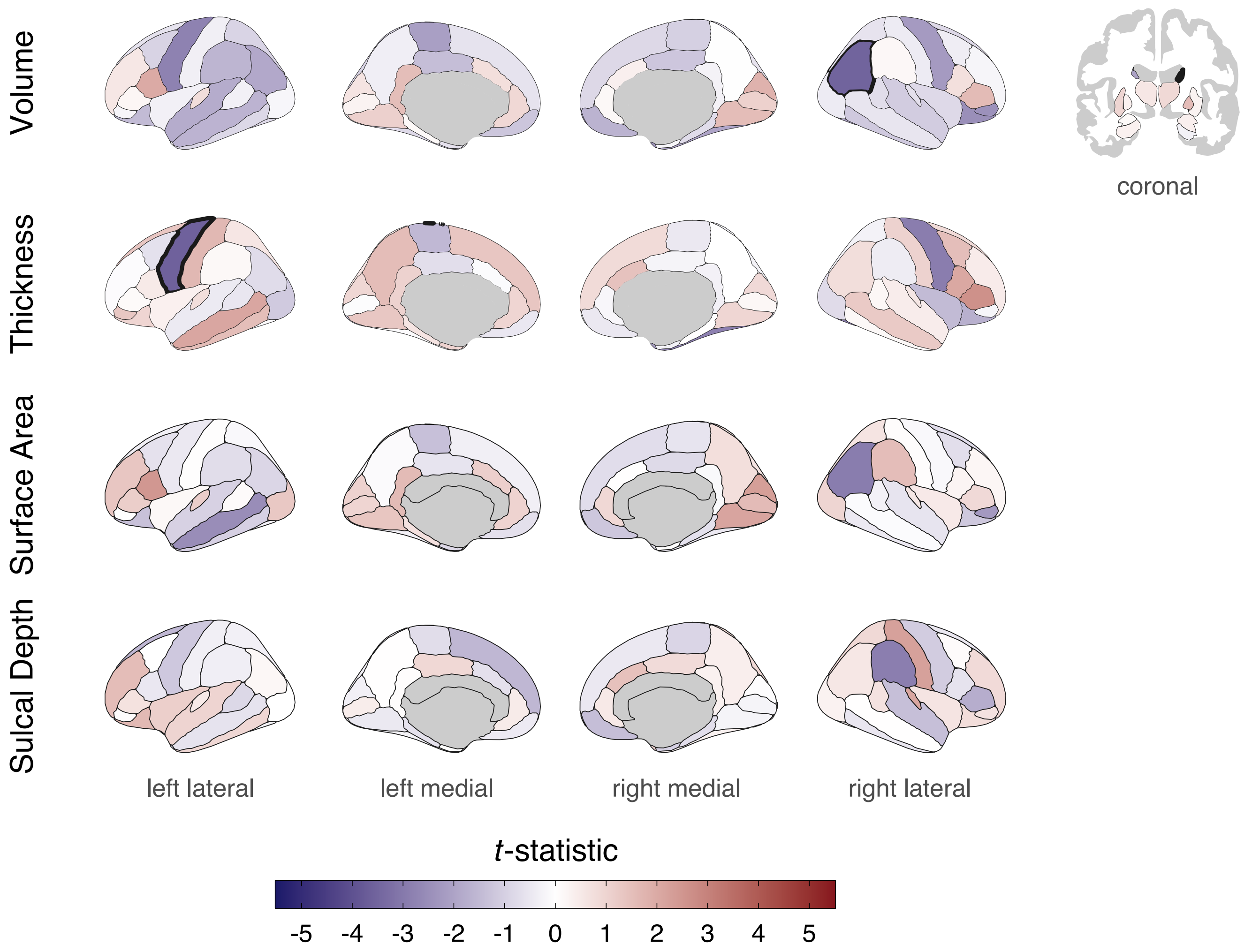


**Figure S4. Regional cortical and subcortical associations with lifetime cannabis use initiation in ABCD.** Cortical and subcortical patterning of associations with substance use plotted as *t*-statistics (red = positive association, blue = negative association). Regions with bold outlines exhibit FDR-significant associations: volume of R inferior parietal and caudate and thickness of L precentral. See also **Table S9.**

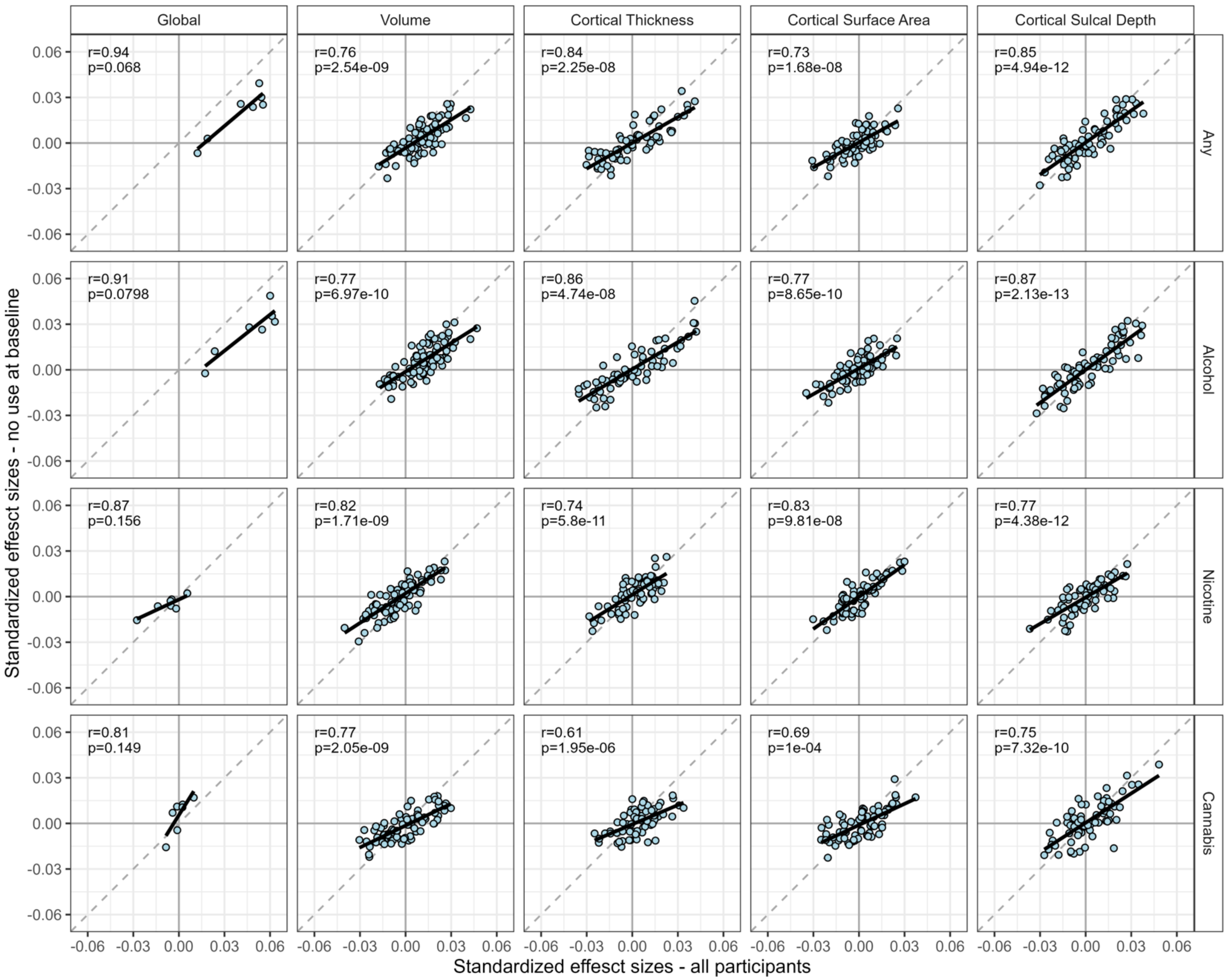


**Figure S5**. **Correlations between effect sizes in analyses in the full sample and analyses restricted to participants without baseline substance use initiation.** Exchangeability block permutations with accelerated *P*-values were used to determine the significance of the correlations. Results show a high correspondence in effect sizes between the two analyses.
